# Unseen but Present: Asymptomatic COVID-19 Cases and Air Travel to Hong Kong

**DOI:** 10.1101/2025.03.26.25324137

**Authors:** Weijun Yu, Hongjie Liu, Deus Bazira, Dylan Ratnarajah, Heran Mane, Thu T. Nguyen, Cheryll Alipio, Xin He, Yulia Hutsul, Jie Chen, Quynh C. Nguyen

**Author notes:** **Corresponding Authors:** Weijun Yu, PhD, MD, MS, Quynh C. Nguyen, PhD MSPH.

## Abstract

The global spread of infectious diseases was influenced by human movement dynamics, particularly for highly transmissible diseases like COVID-19. Asymptomatic COVID-19 cases lacked symptoms before diagnosis, posing a challenge for containment. Their contribution to air travel remains understudied. This retrospective cross-sectional study investigated the role of asymptomatic COVID-19 cases in air travel and their impact on the global spread of the virus. Through our analysis of 11,775 COVID-19 cases in Hong Kong (January 2020–April 2021), log-binomial regression models assessed the association between asymptomatic status and air travel behavior 14 days before diagnosis. The Wilcoxon rank-sum test compared median flight durations between asymptomatic and symptomatic cases. Results revealed two-thirds of cases with air travel history were asymptomatic, with asymptomatic airport or flight crew ten times more likely to travel than symptomatic counterparts (adjusted PRR=10, 95% CI: 4.00–25.00). For non-crew individuals, the adjusted PRR was 1.14 (95% CI: 1.12–1.16). Median flight duration for asymptomatic cases was 4.6 person-hours shorter than symptomatic ones (p<0.01). These findings highlight the significant contribution of asymptomatic cases to air travel and suggest under-detection during initial travel restrictions. Our study emphasizes proactive public health measures early in pandemics involving airborne infections, irrespective of symptom presentation.

## Introduction

In December 2019, the first human-to-human transmissible coronavirus disease (COVID-19), caused by SARS-CoV-2 was reported in Wuhan, China [1]. As the pandemic emerged, research showed that asymptomatic cases could transmit the virus [2, 3]. The World Health Organization (WHO) defines an asymptomatic case as those who test positive for SARS-CoV-2 without showing symptoms [4]. Asymptomatic cases comprise over one-third of diagnosed COVID-19 cases [5]. However, assessing the transmissibility of asymptomatic cases was challenging due to the nature of viral transmission and the difficulty in identifying these cases without widespread testing [6].

Global air travel was intricately linked to infectious diseases [7, 8]. With over three billion passengers traveling annually, the risk of airborne disease transmission, including COVID-19, increased during flights, particularly when passengers were in close proximity to infected individuals [9, 10, 11]. Asian airport hubs were susceptible to the pandemic [12], while some North American and European regions with high Fragile State Indexes (measuring vulnerabilities contributing to global outbreaks) were also vulnerable [13, 14]. In response, various strategies were implemented to reduce in-flight transmission [15], including mask-wearing, sanitized deep cleaning of aircrafts, improved air circulation, pre-flight testing, quarantines, and post-flight contact training [16, 17]. Despite these interventions, in-flight transmission remained possible, as evidenced by six asymptomatic cases on a flight from Milan to South Korea in March 2020 [18]. Thus, identifying asymptomatic cases was critical for managing infectious diseases spread [19].

Asymptomatic transmission has also been observed in outbreaks beyond COVID-19. Asish et al. found that most dengue virus cases during outbreaks were asymptomatic [20], while asymptomatic infections contributed significantly to disease transmission, despite lower viremia levels [21]. Similarly, a review of asymptomatic malaria cases in Southeast Asia revealed differing epidemiological characteristics, complicating public health responses [22]. In South Africa, a study on influenza showed that a quarter of secondary infections originated from asymptomatic cases [23]. Understanding the prevalence of asymptomatic cases is vital for effective public health strategies, especially in dynamic, densely populated areas.

Hong Kong, despite its dense population, reported a low COVID-19 prevalence compared to other developed regions [24]. This success was attributed to its experience with previous outbreaks, such as SARS in 2003, and the early adoption of preventive measures like mask-wearing, handwashing, and social distancing [25, 26, 27, 28]. The Hong Kong Department of Health’s detailed reporting of symptomatic status and travel history of confirmed cases improved contact tracing and case investigation.

The impact of asymptomatic COVID-19 cases on air travel behavior has been understudied. To address the knowledge gap, the objective of our study is to investigate the association between asymptomatic COVID-19 status and air travel behavior before diagnosis. Unlike previous studies limited by small sample sizes or single-flight analyses, our study offers a comprehensive understanding by analyzing two years of data. Our hypotheses are: 1) asymptomatic COVID-19 cases are more likely to travel than symptomatic ones; 2) flight durations differ significantly between asymptomatic and symptomatic cases; and 3) demographic and health factors can modify the association of our interest.

## Methods

This study is a retrospective cross-sectional study that used empirical COVID-19 surveillance data in Hong Kong. The Hong Kong Department of Health publicly released reports containing detailed de-identifiable information for COVID-19 cases since the first case was reported on January 23, 2020. Our study utilized this dataset using each case’s unique tracking ID. We utilized the Locally Weighted Scatterplot Smoothing (LOESS) technique to identify peaks in the daily incidence data of COVID-19 cases in Hong Kong. The LOESS curve allowed us to smooth out random fluctuations and noise in the raw data of reported COVID-19 cases, providing a clear view of the critical peaks of the epidemic curves. Our study included cases reported during these key peaks.

Our data provided information on the age, sex, and asymptomatic or symptomatic status of each COVID-19 case, brief medical histories of the cases, indicating whether they had underlying conditions or good health history, as well as their occupation (e.g., flight attendant, pilot, airport staff), as these factors had implications for exposure to asymptomatic COVID-19 cases. Flight records were also included if the case took an inbound flight 14 days prior to COVID-19 diagnosis. Flight duration data was obtained from airlines’ official websites for historical flight records.

### Human Subjects and Ethics Consideration

This study utilized publicly available data on COVID-19 cases from the Hong Kong Department of Health. Data were de-identified by using unique tracking IDs generated by the Hong Kong Department of Health. These tracking IDs were created to protect the identities of the COVID-19 cases while enabling researchers to follow up on important updates regarding hospitalization, discharge, deceased status, and contact tracing reports without compromising the cases’ identities. As the data was publicly available and de-identified, this study was classified as non-human subject research and was formally waived for ethical approval by the Institutional Review Board at the University of Maryland College Park (IRB number 1882289-1).

### Important Variables

We examined several dependent and independent variables to analyze the association of our interests. The dependent variable of *Inbound air travel history* was determined by COVID-19 cases’ travel history within 14 days prior to diagnosis. Cases were categorized as “1” for those with an inbound air travel history and “0” for those without, with data collected from objective sources such as the immigration department, airport, and airlines. *Flight duration*, another dependent variable, was measured in person-hours. When specific flight numbers and travel dates were available, we obtained the information from the airlines’ official websites. In instances where only the departure geographical locations (e.g., cities) were known, flight duration was estimated using similar historical flight itinerary records by major airlines. When only the departure country was known, we used the median flight duration between the longest and shortest historical itineraries from the country of origin to Hong Kong. Of the cases analyzed, 79.5% had specific flight number and travel date records, 1.6% had records of departure cities, and 18.9% had records of departure countries.

For independent variables, *Asymptomatic status* was determined based on daily situation reports into three categories: asymptomatic (exhibited no symptoms at all), symptomatic (had a date of onset with symptoms), or unknown. *Underlying health conditions* were identified through contact tracing reports and categorized into two groups: cases with known underlying conditions, such as cardiovascular disease or diabetes, and those without underlying conditions, who were previously healthy. *Age* and *Sex* (female or male) were obtained from daily situation reports. *Occupation* was classified based on contact tracing reports into two categories: airport or flight crew (including roles like flight attendant, pilot, and airport staff) and non-airport or flight crew (encompassing all other professions outside the airport or airline industry).

### Potential Bias Assessment

To avoid potential recall bias generated in self-reported data, the Hong Kong Department of Health obtained all reported COVID-19 cases’ travel history for the 14 days prior to diagnosis from objective sources such as the immigration department, airport, and airlines. Measurement bias is unlikely when flight numbers or departure city records are available but is possible when only departure country records exist, as flight duration cannot be exactly determined

### Statistical analysis

Descriptive analysis was conducted on all COVID-19 cases, calculating mean values and standard deviations for age and flight duration as continuous variables. Proportions were calculated for categorical variables, including asymptomatic status, inbound air travel history 14 days prior to diagnosis, sex, occupation, and underlying health condition(s).

To investigate the association between COVID-19 asymptomatic status and inbound air travel history, we utilized log-binomial regression models and reported prevalence risk ratios with 95% confidence intervals. The Hierarchically Well-Formulated Model principle was followed to select the best-fit model. Firstly, all the independent variables, potential effect modifiers, and confounders were included in the full model. Then, the interaction effects of potential modifiers and potential confounders were examined in reduced models. The significance of the interaction terms was determined using the Wald test, and significant effect modifiers were retained in the best-fit model. The 10% rule was applied to determine if variables were confounders by comparing crude and adjusted prevalence risk ratios. A variable was considered a confounder if there was a greater than 10% change between crude and adjusted prevalence risk ratios. The best-fit model retained statistically significant effect modifiers and controlled for confounders. Adjusted prevalence risk ratios were reported by stratification of the significant effect modifier.

To examine differences in flight duration between asymptomatic and symptomatic groups, we conducted a comparison test. First, the normality assumption was tested, and our data violated the normality assumption. Therefore, a nonparametric Wilcoxon rank-sum test was used to compare the two groups

The missing data rate in this study was less than 1%, thus we utilized the pairwise deletion method to manage missing data. A quarter percent (0.25%) of the data (30 out of 11,775 cases) had missing information on cases’ COVID-19 asymptomatic status and sex, and thus were deleted from data analysis. The statistical significance level was set to be 0.05, and a *p*-value smaller than 0.05 was considered statistically significant. The statistical analyses were conducted using SAS OnDemand for Academics (SAS Institute, Cary, NC), and the LOESS curve figure was plotted using the R package ggplot2.

## Results

We included a total of 11,745 COVID-19 cases reported from late January 2020 to April 2021 for analysis. Our study population had a mean age of 44.2 (± 19.3) years old, with over half (n=6,115) being female. Approximately 3% of the cases (n=347) reported underlying health conditions, while 1% (n=104) reported occupations related to airports or airlines. About one-third of the study population (n=3,606) were asymptomatic. Figure 1 presents the temporal pattern of COVID-19 cases in Hong Kong from January 2020 to December 2021. The LOESS curve shows three peaks in the number of daily confirmed COVID-19 cases from late January 2020 to April 2021, followed by a flattening of the curve from May 2021 to December 2021. Therefore, we excluded cases reported after May 2021 from this study.

**Figure 1.**
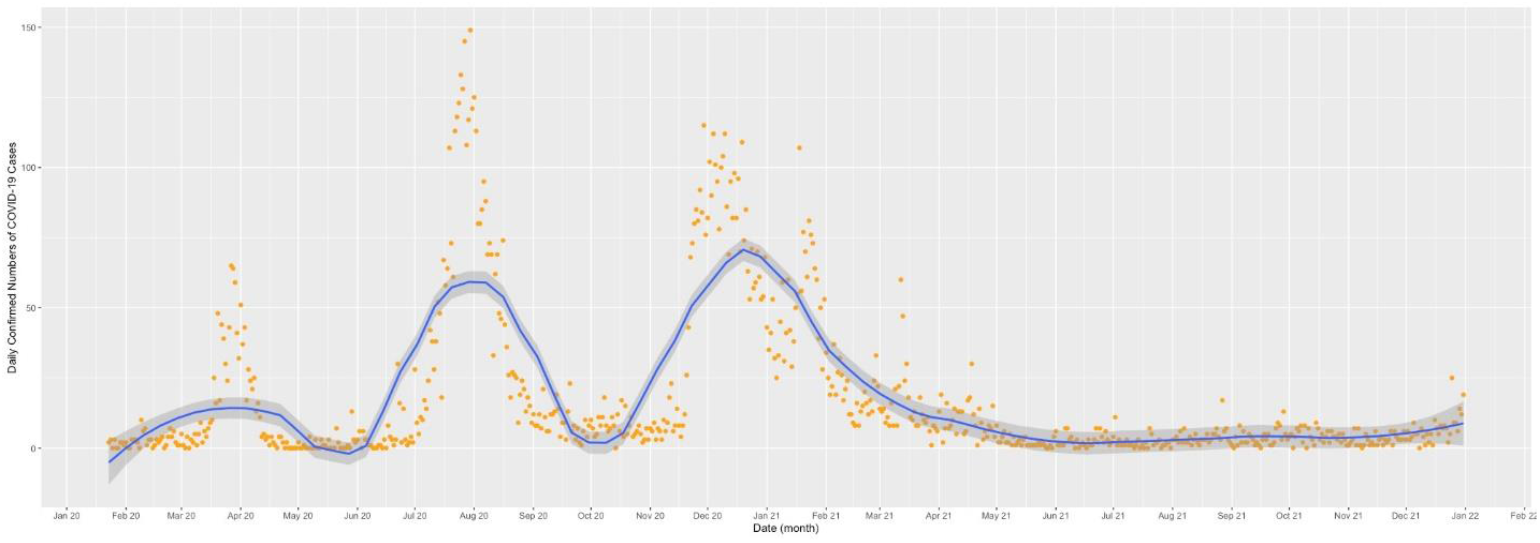
LOESS Curve for COVID-19 Cases in Hong Kong (Jan 2020-Dec 2021)

The Hong Kong government implemented various policies and measures during the three peaks. These measures included border closure to non-Hong Kong residents, mandatory mask-wearing, tightened screening at airports, and social distancing in public places and on public transportation. The descriptive statistics of the study population indicated that symptomatic cases have a slightly younger mean age (40.3 ±19.9 years old) than symptomatic cases (45.9 ±19.5 years old). The sex distribution of asymptomatic cases was similar to symptomatic cases. About two-thirds of the cases who reported inbound air travel history 14 days prior to diagnosis of COVID-19 were asymptomatic, while 17% of the cases who reported underlying health conditions were asymptomatic. Moreover, 73% of the cases who worked as airport or flight crew were asymptomatic.

### Association between COVID-19 Asymptomatic Status and Inbound Air Travel

Our findings using log-binomial regression model analysis demonstrated a significant association between COVID-19 asymptomatic status and inbound air travel behavior, with this association modified by occupation (airport or flight crew versus other occupations) (p<0.0001). Age was found to be a confounder, as evidenced by the change between crude and adjusted prevalence risk ratios (32.9%, greater than 10%). A directed acyclic graph illustrates the significant effect modifiers and confounders for the association of our interest. After adjusting for age as a confounder, the inbound air travel behavior of airport or flight crew members was significantly associated with an asymptomatic status prior to COVID-19 diagnosis (adjusted PRR=10.00, 95% CI: 4.00 – 25.00). This finding suggests that asymptomatic airport or flight crew members were ten times more likely to have engaged in inbound air travel within 14 days before diagnosis compared to their symptomatic counterparts. Among COVID-19 cases who were not part of the airport or flight crew, a similar significant association was observed (adjusted PRR=1.14, 95%CI: 1.12-1.16). This indicated that asymptomatic cases outside the airport or flight crew group were 1.14 times more likely to have an inbound air travel history 14 days prior to diagnosis than symptomatic cases. Furthermore, sex and underlying health condition(s) were not found to be effect modifiers or confounders after evaluating interaction terms and potential confounding effects.

Our analysis revealed that COVID-19 asymptomatic cases with an inbound air travel history 14 days prior to diagnosis had a shorter median flight duration of 6.5 person-hours (25% Q1: 4.0 - 75% Q3:10.4) compared to symptomatic cases, who had a median flight duration of 11.1 person-hours (25% Q1: 5.0 - 75% Q3:11.7). Our normality test (Kolmogorov-Smirnov test, *p*<0.01) indicates that the flight duration data for both asymptomatic and symptomatic cases were not normally distributed. Therefore, we employed a nonparametric Wilcoxon rank-sum test, which demonstrated a significant difference in flight duration between asymptomatic and symptomatic cases (*p* <0.01). Specifically, asymptomatic cases had a median flight duration that was 4.6 person-hours shorter than that of symptomatic cases (*p*<0.01) for those who had inbound air travel history 14 days before COVID-19 diagnosis.

## Discussion

Our findings indicated that asymptomatic cases were more likely to have traveled by air 14 days before a COVID-19 diagnosis than symptomatic cases. This supports our hypothesis that the absence of symptoms encourages air travel, and is crucial for managing airborne infectious diseases, especially early in pandemics when rapid pre-boarding testing is unavailable. Without symptoms, individuals are more likely to continue traveling. Consequently, Health authorities should strongly advise the public to implement effective prevention strategies like wearing masks and social distancing during traveling, regardless of symptoms. Since distinguishing between respiratory infections like influenza or COVID-19 is difficult for the general public, public health measures remain the most efficient way to prevent pandemics.

Our findings also revealed that asymptomatic cases had shorter median flight durations, but this does not imply lower infection risk. First, early pandemic testing focused on symptomatic individuals, resulting in our dataset capturing mostly long-duration flights among symptomatic cases while missing many asymptomatic ones. Secondly, by mid-2020, countries like the United States and the United Kingdom, which are geographically distant from Hong Kong, restricted symptomatic travelers. This shifted our data toward predominantly short-haul flights from closer countries like India and the Philippines. Moreover, symptomatic individuals were generally discouraged from traveling, increasing detection of asymptomatic cases.

The healthy worker effect [29] was evident in our findings, as workers often have a lower mortality rate than the general population due to better overall health. In our study, 73% of flight crew were asymptomatic before diagnosis, compared to 31% of the general study population. This may be attributed to the airline industry’s routine health assessments, which ensure employees meet fitness-to-fly standards, minimizing the risk of health-related incidents mid-flight among the crew. This process may influence hiring decisions, particularly affecting individuals with underlying health conditions. Whether deliberate or inadvertent, baseline health assessments may result in the exclusion of applicants with underlying health conditions, leading to a generally healthier cohort. Only 17% of cases with underlying conditions in our study were asymptomatic, suggesting that healthier individuals were more likely to remain asymptomatic. Asymptomatic crew members were ten times more likely to have traveled before diagnosis than symptomatic crew members, highlighting the need for targeted strategies for this group.

One strength of our study is the use of objective data, like lab results and flight history, minimizing recall bias. This distinguishes our study from others that rely on self-reported data [30]. Hong Kong’s role as an international transport hub allowed for a diverse population, making our findings applicable to other international hubs. Another strength is the heterogeneity of departure cities in our inbound flight data, making the findings more comprehensive and generalizable. Furthermore, the use of ethically de-identified individual-level data to compare asymptomatic and symptomatic cases in air travel adds robustness to our study.

There are limitations to acknowledge. Our study only examined inbound flights to Hong Kong, excluding outbound flights due to follow-up challenges. Among those with inbound flight records, 70% were Hong Kong residents, 10% were non-residents, and 20% had unknown residency status, making the findings most useful for tracking inbound flights. Being an observational study using secondary data, we lacked direct access to COVID-19 cases, limiting our ability to address data gaps, especially regarding underlying conditions.

## Conclusion

The majority of individuals with a travel history before diagnosis were asymptomatic, suggesting that a substantial proportion of travelers may have unknowingly contributed to the spread of COVID-19. Asymptomatic individuals had shorter flight durations than those of symptomatic individuals, suggesting potential under-detection of asymptomatic cases during the initial implementation of air travel restrictions at the onset of the pandemic.

Our findings underscore the importance of effective and comprehensive travel-related public health policies and practices to manage airborne infectious disease outbreaks from the outset. Public health measures should consider the nature and risk of disease transmission during travel, particularly for asymptomatic but infected individuals. Novel surveillance tools, efficient coordination among global stakeholders, and effective risk communication strategies targeting highly mobile individuals and public transportation workers, including frontline air travel staff, are essential.

## Author Contribution

Weijun Yu (Conceptualization [lead], Data curation [equal], Formal analysis [lead], Methodology [lead], Software [lead], Writing—original draft [lead], Writing—review & editing [equal]), Hongjie Liu (Conceptualization [equal], Methodology [equal], Writing—original draft [equal], Writing—review & editing [equal]), Deus Bazira (Writing—original draft [equal], Writing—review & editing [lead]), Heran Mane (Data curation [lead], Formal analysis [equal]), Thu T. Nguyen (Conceptualization [equal], Methodology [equal], Writing—review & editing [equal]), Dylan Ratnarajah (Writing—original draft [equal],Writing—review & editing [equal], visualization [lead]), Cheryll Alipio (Conceptualization [equal], Writing—review & editing [equal]), Xin He (Conceptualization [equal], Methodology [equal]), Yulia Hutsul (Writing—review & editing [equal], visualization [equal]), Jie Chen (Conceptualization [equal], Methodology [equal]), Quynh C. Nguyen (Conceptualization [equal], Methodology [equal], Writing—original draft [equal], Writing—review & editing [equal], supervision [lead], project administration [lead])

## Data Availability

Data is available in the public source and repository.

